# Distinguishing Relapse from Reinfection in Recurrent Tuberculosis: A Genomic and Epidemiologic Study in Brazil

**DOI:** 10.64898/2026.04.07.26350349

**Authors:** Paulo Cesar Pereira dos Santos, Thais Oliveira Goncalves, Eunice Atsuko Totumi Cunha, Katharine S. Walter, Evelyn Lepka de Lima, Julio Croda, Jason R. Andrews, Crhistinne Cavalheiro Maymone Gonçalves, Kesia Esther da Silva

## Abstract

**Background:** Tuberculosis recurrence represents a substantial proportion of incident tuberculosis in many settings. Distinguishing between its mechanisms can inform public health interventions for prevention.

**Methods:** We conducted a retrospective study of individuals with multiple culture-confirmed TB episodes and paired isolates collected between 2012 and 2023 in Dourados and Campo Grande, Mato Grosso do Sul, Brazil. Patients were classified as having recurrent TB after treatment completion or retreatment following non-curative outcomes. Whole-genome sequencing was used to assess pairwise genetic distances between isolates, classifying relapse or persistent infection (≤12 single-nucleotide polymorphisms [SNPs]) versus reinfection or retreatment with reinfection (>12 SNPs).

**Results:** Among 9,293 individuals with TB, 772 recurrent or retreatment episodes were identified. Paired isolates were available for 82 individuals. Among those who completed treatment, reinfection accounted for 74.1% (40/54) of recurrent episodes, while 25.9% (14/54) were relapse. Among individuals with non-curative outcomes, persistent infection (53.6%, 15/28) and retreatment with reinfection (46.4%, 13/28) occurred at similar frequencies. Relapse and persistent infection occurred earlier after the initial episode, whereas reinfection predominated after two years. Incarceration history was strongly associated with reinfection both after treatment completion (92.5%, p=0.012) and non-curative outcomes (76.9%, p=0.016).

**Conclusions:** In this high-burden setting, reinfection is the main driver of TB recurrence after treatment completion, particularly at longer intervals, indicating ongoing transmission. Relapse and persistent infection remain clinically important, especially following non-curative outcomes. These findings underscore the need for integrated strategies combining adherence support to prevent treatment-related recurrence with interventions to reduce transmission, particularly in high-risk settings.

**Summary:** Whole-genome sequencing revealed that tuberculosis recurrence in Brazil is predominantly driven by reinfection, especially after treatment completion and at longer intervals, while relapse and persistent infection occur earlier and are linked to incomplete treatment, highlighting the need for combined treatment and transmission-control strategies.

## Introduction

Tuberculosis (TB) remains one of the leading infectious causes of morbidity and mortality worldwide. In 2025, the World Health Organization estimated 8.3 million individuals were newly diagnosed with TB and 1.23 million deaths, underscoring the persistent global burden of this disease despite extensive control efforts [1]. Although standard multidrug chemotherapy effectively cures the majority of patients, a subset of individuals experience recurrent tuberculosis, a new episode after completing treatment, representing a critical challenge for both clinical care and public health [2–4]. Repeated TB episodes contribute to ongoing transmission, worsens patient outcomes, and undermines global elimination efforts [2,4].

Among individuals who complete treatment, a subsequent episode of TB is generally referred to as recurrent TB and may arise through two distinct mechanisms. Relapse reflects endogenous reactivation of the same strain after incomplete clearance, while reinfection results from the acquisition of a new infection following treatment completion [2,4,5]. Differentiating between these mechanisms is crucial for informing public health strategies, as they have different clinical and epidemiological implications. Relapse points to gaps in treatment effectiveness, adherence, or immunologic clearance, whereas reinfection suggests ongoing transmission and vulnerability to repeated exposure. The relative contributions of relapse and reinfection vary across settings and populations.

Previous studies have reported highly variable recurrence rates, with pooled estimates of 2.26 per 100 person years at risk (95% CI 1.87 – 2.73), ranging from 1.47 per 100 person years in low-incidence settings to 4.10 per 100 person years in high-incidence settings (95% CI 2.67 – 6.28), and with relapses accounting for approximately 70% of recurrent cases overall [3,4]. This heterogeneity in recurrence rates reflects the influence of multiple risk factors, including inadequate or incomplete treatment [2], HIV coinfection [6], diabetes mellitus [7], specific *M. tuberculosis* strain lineages [8], imprisonment [3], alcohol use [9], and smoking [10]. Identifying the factors associated with different recurrence mechanisms is essential for informing targeted interventions and reducing recurrent disease [2–4].

However, repeated TB episodes do not occur exclusively after treatment completion. A distinct but underexplored clinical scenario arises among individuals who do not achieve a curative treatment outcome and later re-present for TB care. In these cases, the subsequent episode may reflect persistent infection with the original strain following incomplete bacterial clearance, or reinfection with a new strain acquired through continued exposure. In contrast to recurrent TB after cure, the relative contribution of these pathways remains poorly characterized. This distinction is clinically and epidemiologically important, as persistent infection and reinfection after treatment non-completion likely require different management and prevention strategies [2,3].

Molecular epidemiology has transformed the study of repeated TB episodes [11].

Genotyping tools like IS6110 RFLP and MIRU-VNTR offered limited resolution, but whole-genome sequencing (WGS) now provides high resolution for distinguishing relapse from reinfection, using robust SNP-distance thresholds and enabling detailed insights into strain diversity, transmission links, and resistance mutations [12,6,13]. Yet most genomic studies have focused primarily on recurrent TB after treatment completion, with less attention to episodes occurring after incomplete treatment. In this study, we integrated WGS with detailed clinical and epidemiological data to investigate the mechanisms underlying repeated TB episodes in two high-burden municipalities in Brazil. Specifically, we examined both recurrent TB after treatment completion and retreatment following incomplete treatment, with the aim of distinguishing relapse, persistent infection, and reinfection, and identifying factors associated with each pathway.

## Methods

### Study population

In this retrospective cohort study, we examined individuals who experienced more than one episode of tuberculosis between 2012 and 2023 in the municipalities of Dourados and Campo Grande, located in the state of Mato Grosso do Sul, Brazil. Cases were identified through the Brazilian Information System for Notifiable Diseases (SINAN). We included individuals with two or more episodes of culture-positive TB for whom *M. tuberculosis* isolates from consecutive episodes were available for whole-genome sequencing (WGS). Sociodemographic and clinical data were obtained from SINAN, and incarceration history was retrieved from the State Incarceration Database (SIGO). All participants provided written informed consent prior to study participation. This study received approval from the Research Ethics Committees of the Federal University of Grande Dourados, the Federal University of Mato Grosso do Sul, the Brazilian National Research Ethics Committee (CONEP) (3.780.597, CAAE 26620619.6.0000.0021; 3.483.377, CAAE 37237814.4.0000.5160; and 3.483.377, CAAE 37237814.4.0000.5160), and the Stanford University Institutional Review Board. Individuals were excluded if the interval between consecutive episodes, defined as the time between sample isolation dates, was less than six months, to reduce misclassification due to prolonged culture positivity or delayed diagnosis. Additional exclusion criteria included failure of subculture, contamination, or evidence of mixed infection, unsuccessful DNA extraction, or sequencing failure.

### Clinical and epidemiological classification

Individuals were classified into two clinical and epidemiological scenarios based on the treatment outcome of their initial TB episode. First, individuals who completed treatment for their initial TB episode and subsequently developed another episode were classified as having recurrent tuberculosis. Treatment completion was defined as a recorded outcome of “cure” in SINAN, an administrative classification based on clinical, radiologic, and/or microbiological criteria, with a treatment duration consistent with standard therapy (5-6 months), based on the interval between treatment initiation and case closure dates. Second, individuals who did not complete treatment for their initial TB episode and subsequently re-presented for TB care were classified as retreatment following a non-curative outcome. This group included individuals with documented outcomes such as treatment abandonment, loss to follow-up, or other non-curative outcomes in SINAN.

### Whole genome sequencing and bioinformatics analyses

We sequenced *M. tuberculosis* genomes from cultured isolates (S1 Text) on an Illumina NovaSeq X Plus system configured for 2 × 150 bp. Genomic variations were analyzed through a publicly available pipeline on GitHub (https://github.com/ksw9/mtb-call2) (S1 Text) [14].

Lineages and indications of mixed infection were inferred using TBProfiler v4.2.0, which also identified mutations associated with drug resistance [15]. Processed variant calls and consensus sequences were used for phylogenetic reconstruction and pairwise genomic comparisons between sequential isolates from the same individual.

### Phylogenetic reconstruction

Maximum-likelihood phylogenetic trees were reconstructed from high-quality SNP alignments generated using SNP-sites [16]. Trees were inferred in IQ-TREE v2.2.0 with model selection performed using ModelFinder and ascertainment bias correction applied for SNP-only alignments [17]. Node support was assessed using 1,000 ultrafast bootstrap replicates. Additional details are provided in S1 Text.

### Genomic analysis and SNP-based classification of recurrence and retreatment

Pairwise single-nucleotide polymorphism (SNP) differences between sequential *M. tuberculosis* isolates from the same individual were calculated in R using the *ape* package [18]. Based on established thresholds [19–21], genomic comparison of paired isolates was used to classify consecutive TB episodes according to strain-relatedness. When more than two sequential isolates were available from the same individual, only the first and second episodes were included in the primary paired analysis.

Among individuals classified clinically as having recurrent tuberculosis, paired isolates differing by ≤12 SNPs were classified as recurrence due to relapse, consistent with reactivation or persistence of the same strain. In contrast, paired isolates differing by >12 SNPs were classified as recurrent due to reinfection, consistent with infection by a genetically distinct strain following treatment completion. Among individuals undergoing retreatment following a non-curative outcome, paired isolates differing by ≤12 SNPs were classified as retreatment with persistent infection, consistent with ongoing infection by the same strain. Paired isolates differing by >12 SNPs in this group were classified as retreatment with reinfection, indicating acquisition of a genetically distinct strain despite a non-curative outcome of the initial episode.

To evaluate the robustness of strain classification, sensitivity analyses were performed using a more stringent threshold of 5 SNP differences to define close genetic relatedness. In addition, we performed minority variant analysis specifically in relapse pairs to investigate whether fixed differences observed between sequential isolates could be explained by the presence of a minority variant in the preceding episode. Minority variants were defined as positions with two or more alleles, each supported by at least 5× coverage, with the minor allele frequency (MAF) above 1%, across the full *M. tuberculosis* genome. For each relapse pair, we evaluated sites fixed in either isolate and tested whether the alternate allele appeared as a minority variant in the corresponding paired sample, thereby assessing minority-allele spillover in both directions.

### Statistical Analyses

Sociodemographic and clinical variables from the first episode were summarized and compared between the relapse and reinfection groups. Categorical variables were evaluated with the chi-squared test, and continuous variables were analyzed with the Wilcoxon rank-sum test. In addition, pairwise SNP distances between sequential isolates were modeled as a function of the time interval between episodes to explore temporal dynamics of divergence. All analyses were performed using R v4.3.0.

## Results

### Classification of recurrence: relapse vs reinfection

Between 2012 and 2023, a total of 9,293 individuals with TB were reported in Dourados and Campo Grande. Of these, cultures were not performed for 4,286 patients, and 1,407 had negative cultures, leaving 3,600 culture-positive patients. Among these, 772 individuals experienced a subsequent TB episode (Figure 1). We excluded 179 recurrences or retreatment episodes occurring within six months of the prior episode, using sample isolation dates to define intervals between episodes, resulting in 593 episodes occurring more than six months apart.

**Figure 1.**
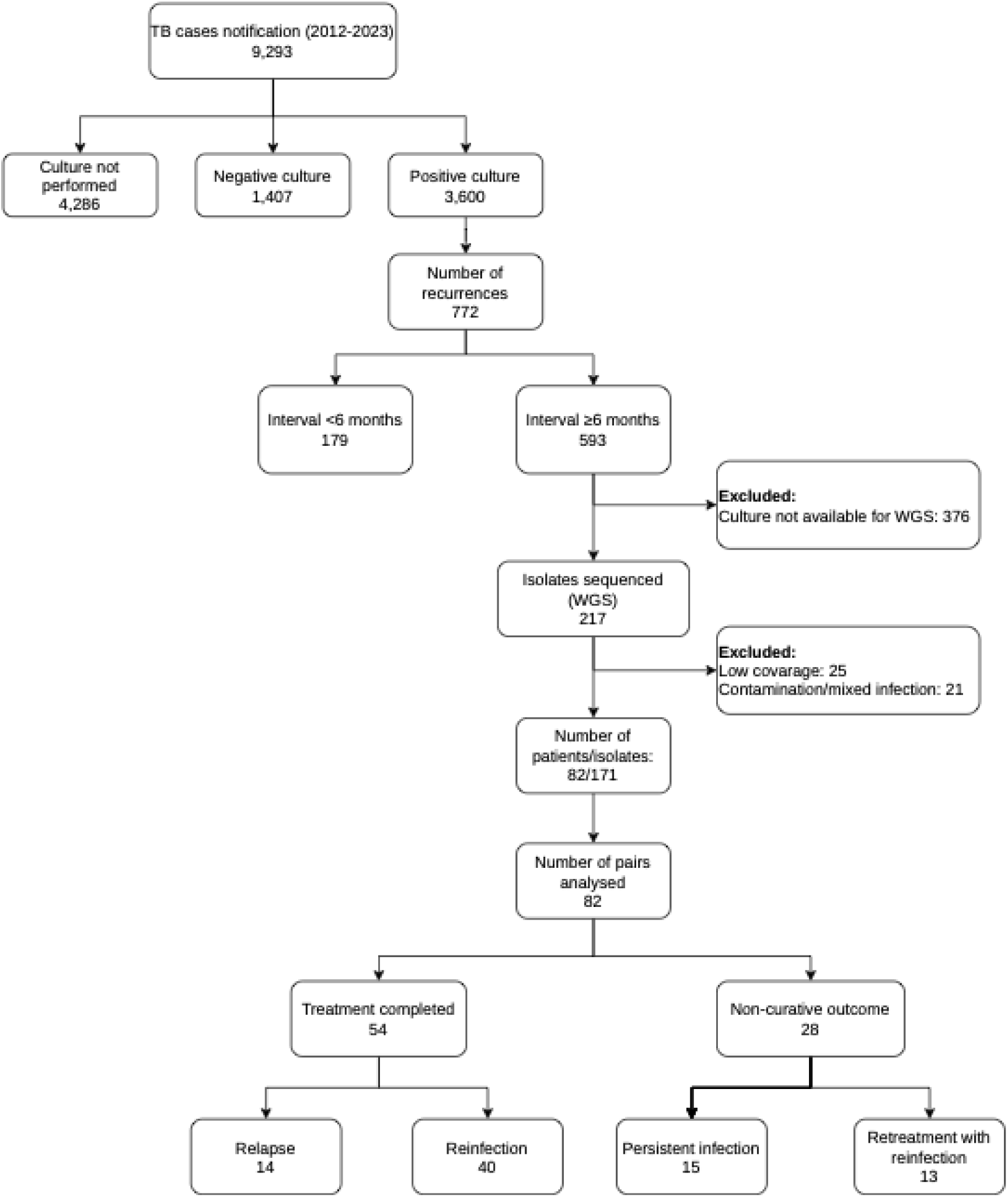
Diagram with the discarded and selected individuals for the recurrent cases study. WGS denotes whole-genome sequencing.

From this group, cultures were unavailable for 376 individuals, leaving 217 isolates eligible for whole-genome sequencing (WGS). Following sequencing, 46 isolates were excluded due to low coverage (n = 25) or evidence of contamination and/or mixed infection (n = 21). The final dataset comprised 171 high-quality isolates from 82 individuals, generating 82 paired isolate comparisons. Most individuals experienced two TB episodes (76/82, 92.7%), while five (6.1%) experienced three episodes and one (1.2%) experienced four episodes. For individuals with more than two episodes, the analysis was restricted to isolates from the first two episodes and their corresponding outcomes.

Pairwise SNP comparisons enabled classification of subsequent episodes according to clinical context and genomic relatedness. Overall, 54 individuals (65.8%) completed treatment for their initial episode. Among these, 14 (25.9%) were classified as recurrence due to relapse (≤12 SNPs between sequential isolates), whereas 40 (74.1%) were classified as recurrence due to reinfection (>12 SNPs). The remaining 28 individuals (34.1%) did not complete treatment.

Within this group, 14 (53.6%) were classified as persistent infection with the same strain (≤12 SNPs), and 13 (46.4%) were classified as retreatment with reinfection (>12 SNPs).

### Temporal patterns of relapse, reinfection, and retreatment

Among individuals who completed treatment, the timing of relapse and reinfection episodes differed substantially (Figure 2A). Relapse episodes were distributed across the follow-up period, with a median interval of 679 days between episodes (IQR 409–1,285), whereas reinfection episodes occurred later, with a median interval of 1,214 days (IQR 844–1,716; Wilcoxon rank-sum p = 0.034). Half of the relapse episodes (7/14, 50.0%) occurred within two years of the prior episode, with the remainder occurring after two years. In contrast, reinfection episodes occurred predominantly after two years (34/40, 85.0%), with only six (15.0%) occurring within the first two years (Fisher’s exact p < 0.025). Among relapse pairs, we did not identify a relationship between time interval and SNP differences (Figure 2C).

**Figure 2.**
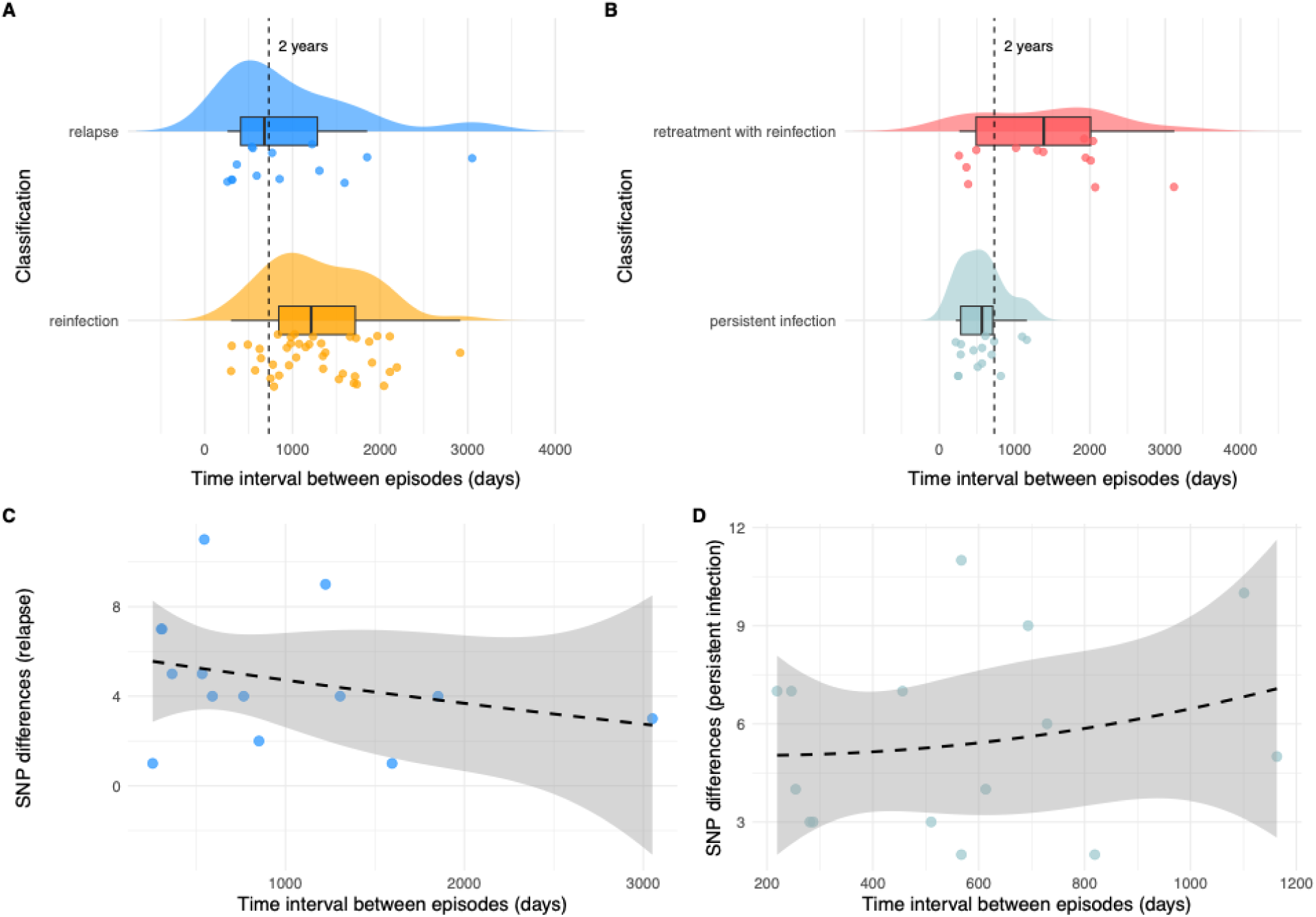
Temporal patterns and genomic relatedness of sequential *Mycobacterium tuberculosis* isolates. (**A**) Distribution of time intervals between sequential tuberculosis episodes among individuals who completed treatment, classified as recurrence due to relapse (≤12 SNP differences) or reinfection (>12 SNP differences). (**B**) Distribution of time intervals between sequential episodes among individuals undergoing retreatment following a non-curative outcome, classified as persistent infection (≤12 SNP differences) or retreatment with reinfection (>12 SNP differences). In panels A and B, raincloud plots display kernel density estimates (half violins), boxplots indicating median and interquartile range, and jittered points representing individual episode pairs. The vertical dashed line denotes a 2-year interval between episodes. (**C**) Relationship between the time interval separating episodes and pairwise SNP differences among relapse pairs. (**D**) Relationship between the time interval separating episodes and pairwise SNP differences among persistent infection pairs. Each point in panels C and D represents a pair of *M. tuberculosis* isolates obtained from the same individual during consecutive TB episodes.

Distinct temporal patterns were also observed among individuals undergoing retreatment following a non-curative outcome in the first TB episode (Figure 2B). Episodes classified as persistent infection occurred earlier, with a median interval of 567 days (IQR 284–711), compared with retreatment with reinfection, which occurred later (median 1,388 days, IQR 489–2,009; Wilcoxon rank-sum p = 0.013). Persistent infection accounted for most episodes occurring within two years (10/15, 66.7%), whereas retreatment with reinfection was more frequently observed after two years (9/13, 69.2%), although this difference did not reach statistical significance (Fisher’s exact p = 0.130). No relationship was detected between the time interval separating episodes and SNP differences among persistent infection pairs (Figure 2D).

Sensitivity analyses applying a more stringent 5-SNP threshold yielded temporal patterns consistent with the primary analysis (Table S1), indicating that observed timing differences between relapse or persistent infection and reinfection were robust to variation in SNP thresholds.

### Clinical and epidemiological characteristics associated with recurrence patterns

We compared demographic and clinical characteristics recorded at the initial TB episode among individuals who subsequently developed relapse or reinfection (Table 1). Median age did not differ between groups (relapse: 25 years, IQR 24 – 31; and reinfection: 27 years, IQR 23 – 35; p = 0.464). Most individuals were male (94.4%), with a higher proportion in the reinfection group than in the relapse group (100.0% vs. 78.6%, p = 0.020). A history of incarceration was common overall (85.2%) and was more frequent among individuals with reinfection than relapse (92.5% vs. 64.3%; p = 0.012).

**Table 1.** Demographic and clinical characteristics of the initial episode among individuals with TB relapse and reinfection after cure from the initial episode.

| **Variables** | **Total**<br>**n (%)**<br>**54** | **Reinfection**<br>**n (%)**<br>**40 (74.1)** | **Relapse**<br>**n (%)**<br>**14 (25.9)** | **p**<br>**value\*** |
| --- | --- | --- | --- | --- |
| **Sex** |  |  |  | 0.020 |
| **Female** | 3 (5.6) | 0 (0.0) | 3 (21.4) |  |
| **Male** | 51 (94.4) | 40 (100.0) | 11 (78.6) |  |
| **Age, median (IQR)** | 26 (23 – 33) | 27 (23 – 35) | 25 (24 – 36) | 0.464 |
| **Ethnicity** |  |  |  | 0.746 |
| **Asian** | 1 (1.9) | 1 (2.5) | 0 (0.0) |  |
| **Black** | 5 (9.3) | 3 (7.5) | 2 (14.3) |  |
| **Indigenous** | 3 (5.6) | 1 (2.5) | 2 (14.3) |  |
| **Mixed** | 17 (31.5) | 15 (37.5) | 2 (14.3) |  |
| **White** | 17 (31.5) | 10 (25.0) | 7 (50.0) |  |
| **Schooling** |  |  |  | 0.180 |
| **<8 years of schooling** | 33 (61.1) | 24 (60.0) | 9 (64.3) |  |
| **Incarceration status** |  |  |  | 0.012 |
| **Incarcerated** | 46 (85.2) | 37 (92.5) | 9 (64.3) |  |
| **Chest radiography†** |  |  |  | - |
| **Abnormal** | 19 (100.0) | 14 (100.0) | 5 (100.0) |  |
| **HIV status** |  |  |  | 0.834 |
| **Positive** | 1 (1.9) | 1 (2.5) | 0 (0.0) |  |
| **Diabetes** |  |  |  | 0.804 |
| **Yes** | 1 (1.9) | 1 (2.5) | 0 (0.0) |  |
| **Any alcohol use** |  |  |  | 0.856 |
| **Yes** | 6 (11.1) | 5 (12.5) | 1 (7.1) |  |
| **Smoking** |  |  |  | 0.162 |
| **Yes** | 12 (22.2) | 10 (25.0) | 2 (14.3) |  |
| **Drug resistance** |  |  |  | 1.000 |
| **Sensitive** | 52 (96.3) | 39 (97.5) | 13 (92.9) |  |
| **HR-TB** | 2 (3.7) | 1 (2.5) | 1 (7.1) |  |
| **Time of recurrent episode** |  |  |  | 0.023 |
| **Less than 2 years** | 13 (24.1) | 6 (15.0) | 7 (50.0) |  |
| **More than 2 years** | 41 (75.9) | 34 (85.0) | 7 (50.0) |  |

\* p-values generated using the Chi-square test and the Wilcoxon test.
† Percent abnormal includes only participants with non-missing chest radiography results.

Among individuals undergoing retreatment following a non-curative outcome in the first TB episode, demographic and clinical characteristics at the initial episode were compared between those classified as persistent infection and retreatment with reinfection (Table 2).

**Table 2.**
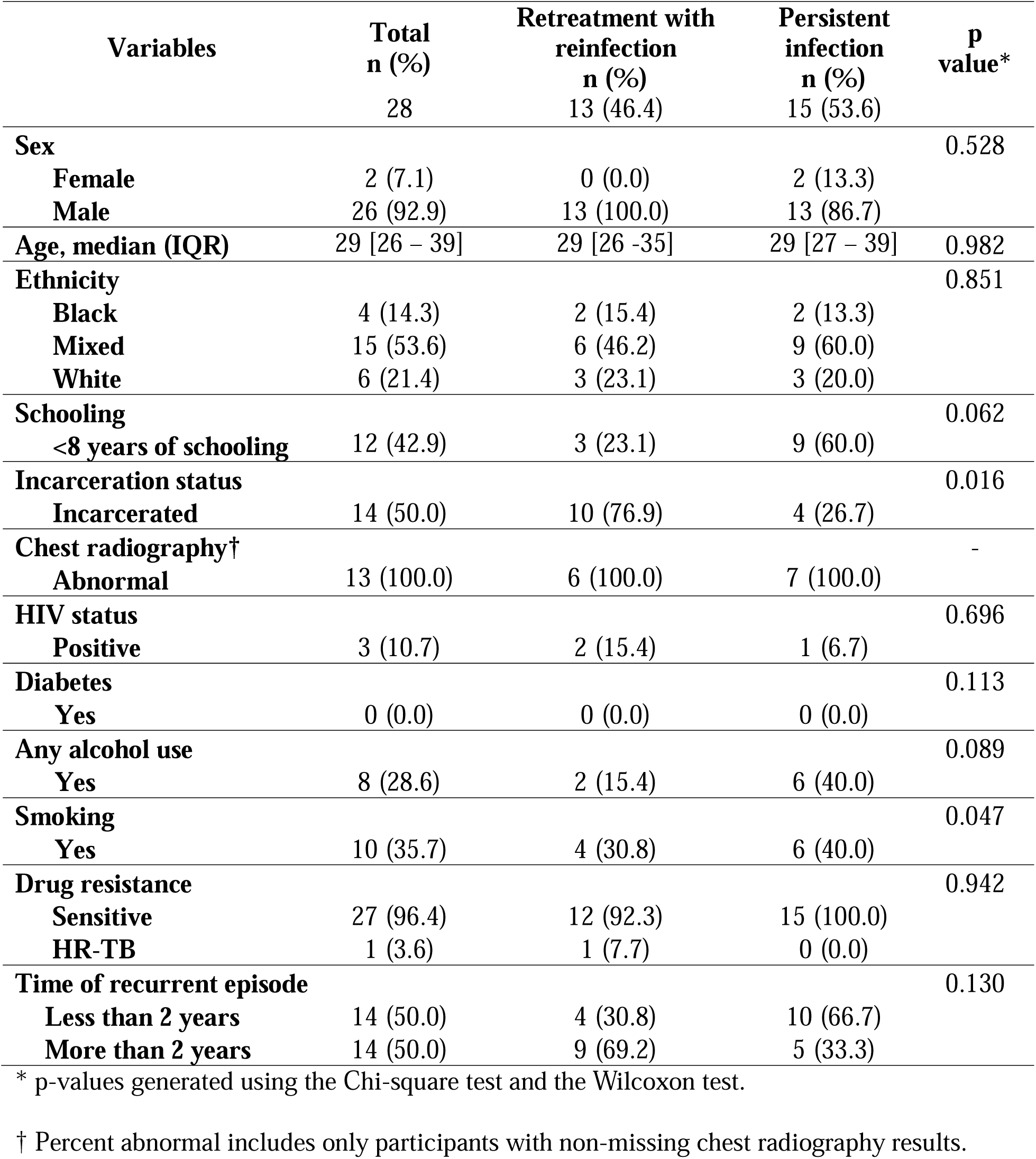
Demographic and clinical characteristics of the initial episode among individuals with persistent TB infection and retreatment with TB reinfection after a non-curative outcome from the initial episode.

Median age was comparable between groups (persistent infection: 29 years, IQR 27 –39; retreatment with reinfection: 29 years, IQR 26–35; p = 0.982), and most participants were male (92.9%) with no significant difference by recurrence classification (p = 0.528). HIV status, diabetes prevalence, and alcohol use did not differ significantly. Smoking status differed, with a higher proportion of non-smokers among individuals with persistent infection (53.3% vs. 23.1%; p = 0.047). Incarceration history was more frequent among individuals with retreatment with reinfection than persistent infection (76.9% vs. 26.7%; p = 0.016).

### Phylogenetic structure of recurrent isolates

Phylogenetic reconstruction showed that all 171 *M. tuberculosis* isolates belonged to Lineage 4 (Figure 3). The most common sub-lineage was 4.1.2.1, accounting for 33.3% (57/171) of the isolates, followed by 4.4.1.1 (21.6%; 37/171), 4.3.3 (17.5%; 30/171), 4.4.1.2 (15.2%; 26/171), and 4.3.4.2 (6.4%; 11/171). The majority of isolates (93.6%, 160/171) were predicted to be drug-sensitive based on genomic markers. Isoniazid monoresistance (HR-TB) was identified in 5.8% (10/171) of the isolates. Furthermore, one isolate (0.6%) exhibited mutations consistent with multidrug-resistant tuberculosis (MDR-TB), and another (0.6%) exhibited genetic alterations associated with fluoroquinolone resistance. Notably, the single MDR-TB isolate was identified in the second episode of a patient classified as retreatment with reinfection, consistent with acquisition of a drug-resistant strain rather than persistence of the original infection. In all pairs in which isoniazid resistance-associated mutations were newly detected, including relapse and persistent infection pairs, these mutations were present only in the second isolate, consistent with resistance emerging or being selected after the initial episode.

**Figure 3.**
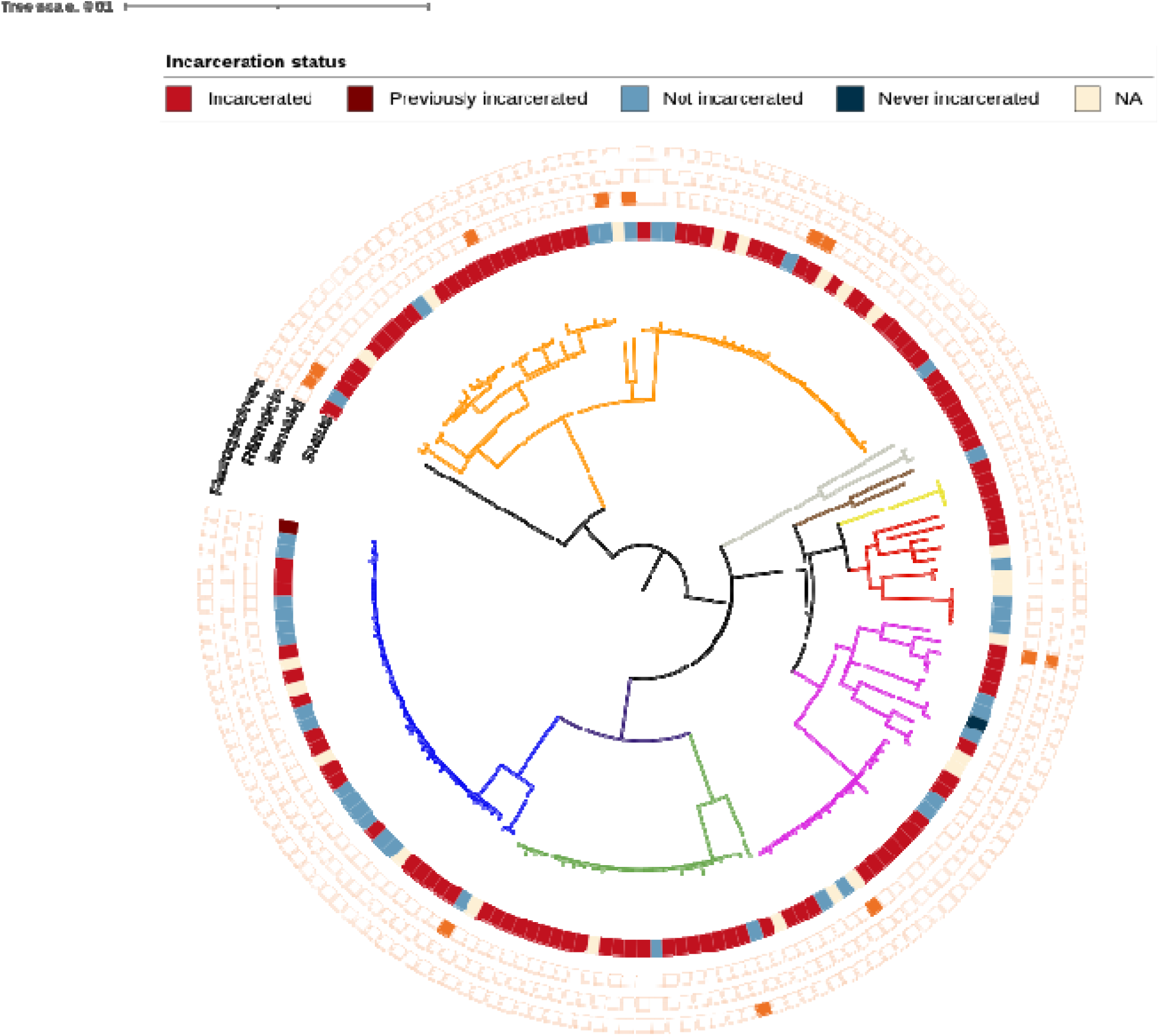
Phylogenetic structure and drug resistance profile of *Mycobacterium tuberculosis* strains from recurrent tuberculosis. A maximum likelihood phylogenetic tree of 171 *Mycobacterium tuberculosis*, obtained from 82 patients with recurrent tuberculosis. Branch lengths represent genetic distance in units of substitutions per site, and branches are colored according to sub-lineage classification. From the center outward, the concentric rings indicate: (1) sub-lineage assignment, (2) incarceration status at the time of TB notification, and (3) antimicrobial resistance prediction. The scale bar indicates the nucleotide substitution per site.

Relapse pairs clustered closely in the phylogeny, while reinfection pairs were distributed across phylogenetically distant branches, consistent with the acquisition of new strains. (Figure S1). The phylogenetic analysis showed that isolates from incarcerated individuals were dispersed across the tree and frequently interspersed with those from community cases (Figure 3), highlighting ongoing transmission between prison and community settings. To evaluate whether genomic differences observed between paired recurrent isolates could be explained by low-frequency variants present at the time of the earlier episode that later became fixed, we evaluated all positions fixed in either isolate or assessed whether the corresponding alternate allele appeared at minority frequency in the partner sample. Across all 31 pairs, no allele fixed in one isolate was detected as a minority variant in its paired isolate. When present in the paired isolate, the same allele was typically near-fixed (AF ≥0.95). Overall, these data indicate low shared within-host heterogeneity between episodes.

## Discussion

This study integrates genomic, epidemiological, and clinical data to investigate the causes underlying multiple tuberculosis episodes among individuals in two high-burden Brazilian municipalities over more than a decade. By combining whole-genome sequencing with patient-level information, we were able to differentiate relapse, reinfection, persistent infection, and retreatment with reinfection with high resolution, revealing distinct temporal and epidemiological patterns across these pathways. This approach allowed us to evaluate recurrence mechanisms among both individuals who completed treatment and those with a non-curative outcome from their initial episode.

Among individuals who completed treatment, reinfection was the predominant mechanism of recurrence, consistent with ongoing exposure in a high-transmission setting. In contrast, relapse accounted for a smaller proportion and showed no clear temporal clustering, occurring both early and late after treatment completion. Among individuals with a non-curative outcome from their initial episode, recurrence was classified as persistent infection or retreatment with reinfection, highlighting that treatment non-completion does not uniformly result in individuals re-presenting with the same dominant strain. Instead, a non-curative outcome appears to give rise to two broad pathways: early recurrence associated with persistence of a closely related strain and later recurrence associated with recovery of a genetically distinct strain. Although these later episodes are most consistent with reinfection, we cannot exclude the possibility that, in some cases, they reflect outgrowth of a previously undetected co-infecting strain present during the earlier episode.

Our findings align with studies from other high-burden settings, where reinfection has been shown to account for a substantial fraction of recurrent tuberculosis, in contrast to low-incidence settings where relapse predominates [22,23]. Reported reinfection proportions ranging from approximately 51% to over 77% in settings such as South Africa [24,25] are comparable to those observed in our cohort. Together, these observations reinforce the notion that treatment success alone is insufficient to prevent future disease in endemic contexts, where sustained transmission continues to place individuals at risk long after successful treatment. In parallel, there is a substantial but less well-characterized burden of retreatment following treatment non-completion in high-burden settings. In Brazil, retreatment after loss to follow-up accounts for nearly half of retreatment cases and is associated with poorer outcomes [26], with similar patterns reported in Kenya and Pakistan [27,28]. However, the mechanisms underlying these episodes remain poorly defined. Our findings extend this evidence by demonstrating that both persistent infection and reinfection contribute to retreatment and are shaped by distinct temporal and epidemiological factors.

The temporal separation between recurrence mechanisms provides further insight into underlying processes. Persistent infection and relapse tended to occur earlier, whereas reinfection and retreatment with reinfection were more frequently observed after longer intervals. This pattern suggests that early recurrence is more closely linked to treatment-related factors, while later recurrence reflects cumulative exposure to circulating strains. Notably, the absence of increasing SNP divergence over time among relapse and persistent infection pairs, coupled with the lack of shared minority variants, indicates that these episodes were driven by reactivation of genetically stable populations.

Incarceration emerged as a key determinant of reinfection and retreatment with reinfection across both clinical contexts. The high proportion of reinfection observed in this study also reflects, in part, the oversampling of incarcerated individuals due to intensive case finding and genomic surveillance conducted in prison settings during the study period. Although our study population was predominantly male, reflecting the demographics of incarceration in Mato Grosso do Sul, the consistent association between incarceration and reinfection highlights structural drivers of risk that extend beyond individual-level behaviors. These findings add to a growing body of evidence demonstrating intense bidirectional transmission between prison and community settings and emphasize the need for targeted interventions in carceral environments [29,30].

Treatment non-completion was strongly associated with early recurrence driven by persistent infection, reinforcing prior evidence that inadequate therapy increases the risk of ongoing disease and early re-presentation [2,4,31]. The observation that reinfection also occurred frequently among individuals with a non-curative outcome, particularly at later time points, further illustrates that treatment interruption may not only fail to clear infection but may also increase vulnerability to subsequent exposure. Differences in smoking status between persistent infection and reinfection groups suggest an additional behavioral factor that may influence recurrence pathways, although these associations warrant cautious interpretation.

Our findings should be interpreted within the context of the study’s limitations. First, the oversampling of incarcerated individuals, who have a much higher risk of tuberculosis exposure, likely resulted in a greater contribution of reinfections, and these results likely do not generalize to the broader population in Brazil. Second, nearly half of the recurrent isolates could not be analyzed due to missing or low-quality culture and sequencing data, which may introduce selection bias. Next, although we employed a SNP threshold-based approach for case classification, there is no universally accepted cutoff, and the potential for misclassification persists. However, the consistency of temporal patterns across thresholds strengthens the robustness of our conclusions. Finally, incomplete data on incarceration history and behavioral factors may have led to misclassification error, likely biasing associations towards the null.

Taken together, our findings highlight that recurrent tuberculosis in high-burden settings arises through multiple pathways shaped by both treatment-related factors and ongoing transmission. Interventions that go beyond treatment success are needed to reduce reinfection, including strategies to lower exposure to circulating strains in high-risk environments. At the same time, relapse and persistent infection remain clinically important, particularly following a non-curative outcome from the initial episode, underscoring the importance of adherence support and patient-centered interventions. Overall, our findings indicate that addressing both effective treatment and sustained transmission control will be essential to reduce recurrent disease and accelerate progress toward TB elimination.

## Acknowledgments

We thank the State Agency of Administration Prisons (AGEPEN) for their full support during the study period, the study participants for their kind cooperation during the data collection process, and the Central Laboratory (LACEN) of the state of Mato Grosso do Sul for the support in the accomplishment of the laboratory tests.

## Data availability

Raw sequence data generated in this study have been deposited under the study accession number PRJEB96387.

## Funding

This work was supported by funding from the U.S. National Institutes of Health (R01 AI130058 and R01 AI149620) and the Brazilian National Research Council (CNPq).

## Competing interests

The authors declare that they have no competing interests.

## Notes

### Competing Interest Statement

The authors have declared no competing interest.

### Author Declarations

This study received approval from the Research Ethics Committees of the Federal University of Grande Dourados, the Federal University of Mato Grosso do Sul, the Brazilian National Research Ethics Committee (CONEP) (3.780.597, CAAE 26620619.6.0000.0021; 3.483.377, CAAE 37237814.4.0000.5160; and 3.483.377, CAAE 37237814.4.0000.5160), and the Stanford University Institutional Review Board.

