## Supplemental material for "Distinguishing Relapse from Reinfection in Recurrent Tuberculosis: A Genomic and Epidemiologic Study in Brazil"

The supplementary material includes supplementary text (S1 Text, Supplementary Methods), Supplementary Table S1, Supplementary Figure S1, and Supplementary References.

S1 Text. Supplementary Methods 2

Supplementary Table S1 4

Supplementary Figure S1 6

Supplementary References 7

### S1 Text. Supplementary Methods

#### Laboratory diagnosis and culture

Sputum samples were cultured using the Ogawa-Kudoh method [1]. The cultures were incubated at 37 °C and observed weekly for up to 60 days. DNA was extracted from *Mycobacterium tuberculosis* isolates using the cetyltrimethylammonium bromide method [2].

#### Whole genome sequencing and bioinformatics analyses

Whole-genome sequencing was performed using the Illumina NovaSeq X Plus system configured for 2 × 150 bp. Genomic variations were analyzed through a publicly available pipeline on GitHub (https://github.com/ksw9/mtb-call2) [3]. In brief, bases with a Phred score under 20 were trimmed, and adapters were removed using Trim Galore v0.6.5 with a stringency level of 3. Additional filtering was done with CutAdapt v4.2, using parameters –nextseq-trim=20, –minimum-length=20, and –pair-filter=any [4]. To minimize contamination, reads were taxonomically classified with Kraken2 [5], retaining only those identified as *Mycobacterium genus*, specifically *M. tuberculosis*. The reads were aligned to the *M. tuberculosis* H37Rv reference genome (NCBI Accession: NC_000962.3) using BWA v0.7.15 [6], and duplicate reads were removed with Sambamba [7]. Variant calling was conducted using GATK v4.1 with HaplotypeCaller set to sample ploidy=1 and GenotypeGVCFs [8]. Variants were retained if they had a minimum depth of 10× and a quality score of at least 40, with non-variant sites included in the final VCF files. Consensus sequences were created using bcftools consensus [9], excluding indels. Single-nucleotide polymorphisms (SNPs) located in repetitive regions, such as PPE and PE-PGRS genes, phages, insertion sequences, and repeats longer than 50 bp, were excluded from further analyses [10]. Lineages and indications of mixed infection were inferred using TBProfiler v4.2.0, which also identified mutations associated with drug resistance. [11]. Processed variant calls and consensus sequences were used for phylogenetic reconstruction and pairwise genomic comparisons between sequential isolates from the same individual.

**Phylogenetic reconstruction**

Phylogenetic reconstruction was conducted by generating full-length consensus FASTA sequences from VCF files, followed by the extraction of a multiple alignment of internal variant sites using SNP-sites [12]. For phylogeny construction, the optimal substitution model was determined using ModelFinder, integrated within IQ-TREE v2.2.0, which evaluated all models incorporating ascertainment bias correction suitable for the SNP-only alignments [13]. The model identified as the best fit, according to the Bayesian Information Criterion (BIC), was TVM+F+ASC+R7. Subsequently, a maximum likelihood (ML) phylogenetic tree was inferred using IQ-TREE, employing 1,000 ultrafast bootstrap replicates to evaluate node support. Phylogenetic trees were visualized using iTOL (Interactive Tree of Life) [14].

**Table S1. Demographic and clinical characteristics of the initial episode among individuals with complete TB treatment and non-curative outcome using a more stringent 5-SNP threshold.**

| Variable | Complete treatment | | | | Non-curative outcome | | | |
|---|---|---|---|---|---|---|---|---|
| | Overall n (%) | Reinfection n (%) | Relapse n (%) | p value* | Overall n (%) | Retreatment with Reinfection n (%) | Persistent infection n (%) | p value* |
| | 54 | 44 (81.5) | 10 (18.5) | | 28 | 20 (71.4 | 8 (28.6) | |
| **Sex** | | | | 0.149 | | | | 1.000 |
| **Female** | 3 (5.6) | 1 (2.3) | 2 (20.0) | | 2 (7.1) | 1 (5.0) | 1 (12.5) | |
| **Male** | 51 (94.4) | 43 (97.7) | 8 (80.0) | | 26 (92.9) | 19 (95.0) | 7 (87.5) | |
| **Age, median (IQR)** | 26 (23 – 33) | 26 (23 - 35) | 25 (24 - 31) | 0.468 | 29 (26 – 39) | 29 (27 – 39) | 27 (26 – 35) | 0.414 |
| **Ethnicity** | | | | 0.257 | | | | 0.985 |
| **Asian** | 1 (1.9) | 1 (2.3) | 0 (0.0) | | | | | |
| **Black** | 5 (9.3) | 3 (6.8) | 2 (20.0) | | 4 (14.3) | 3 (15.0) | 1 (12.5) | |
| **Indigenous** | 3 (5.6) | 2 (4.5) | 1 (10.0) | | | | | |
| **Mixed** | 17 (31.5) | 15 (34.1) | 2 (20.0) | | 15 (53.6) | 11 (55.0) | 4 (50.0) | |
| **White** | 17 (31.5) | 12 (27.3) | 5 (50.0) | | 6 (21.4) | 4 (20.0) | 2 (25.0) | |
| **Schooling** | | | | 0.072 | | | | 0.378 |
| **<8 years of schooling** | 33 (61.1) | 27 (61.4) | 6 (60.0) | | 12 (42.9) | 7 (35.0) | 5 (62.5) | |
| **Incarceration status** | | | | 0.191 | | | | 0.913 |
| **Incarcerated** | 46 (85.2) | 39 (88.6) | 7 (70.0) | | 14 (50.0) | 10 (50.0) | 4 (50.0) | |
| **Chest radiography†** | | | | | | | | - |
| **Abnormal** | | | | | 13 (100.0) | 8 (100.0) | 5 (100.0) | |
| **HIV status** | | | | 0.644 | | | | 0.956 |
| **Positive** | 1 (1.9) | 1 (2.3) | 0 (0.0) | | 3 (10.7) | 2 (10.0) | 1 (12.5) | |

| | | | | | | | | |
|---|---|---|---|---|---|---|---|---|
| **Diabetes** | | | | 0.714 | | | | 0.827 |
| **Yes** | 1 (1.9) | 1 (2.3) | 0 (0.0) | | 0 (0.0) | 0 (0.0) | 0 (0.0) | |
| **Any alcohol use** | | | | 0.938 | | | | 0.694 |
| **Yes** | 6 (11.1) | 5 (11.4) | 1 (10.0) | | 8 (28.6) | 5 (25.0) | 3 (37.5) | |
| **Smoking** | | | | 0.426 | | | | 0.509 |
| **Yes** | 12 (22.2) | 10 (22.7) | 2 (20.0) | | 10 (35.7) | 6 (30.0) | 4 (50.0) | |
| **Drug resistance** | | | | 1.000 | | | | 1.000 |
| **Sensitive** | 52 (96.3) | 42 (95.5) | 10 (100.0) | | 27 (96.4) | 19 (95.0) | 8 (100.0) | |
| **HR-TB** | 2 (3.7) | 2 (4.5) | 0 (0.0) | | 1 (3.6) | 1 (5.0) | 0 (0.0) | |
| **Time of recurrent episode** | | | | 0.317 | | | | 0.209 |
| **Less than 2 years** | 13 (24.1) | 9 (20.5) | 4 (40.0) | | 14 (50.0) | 8 (40.0) | 6 (75.0) | |
| **More than 2 years** | 41 (75.9) | 35 (79.5) | 6 (60.0) | | 14 (50.0) | 12 (60.0) | 2 (25.0) | |

* p-values generated using the Chi-square test and the Wilcoxon test.

† Percent abnormal includes only participants with non-missing chest radiography results

.

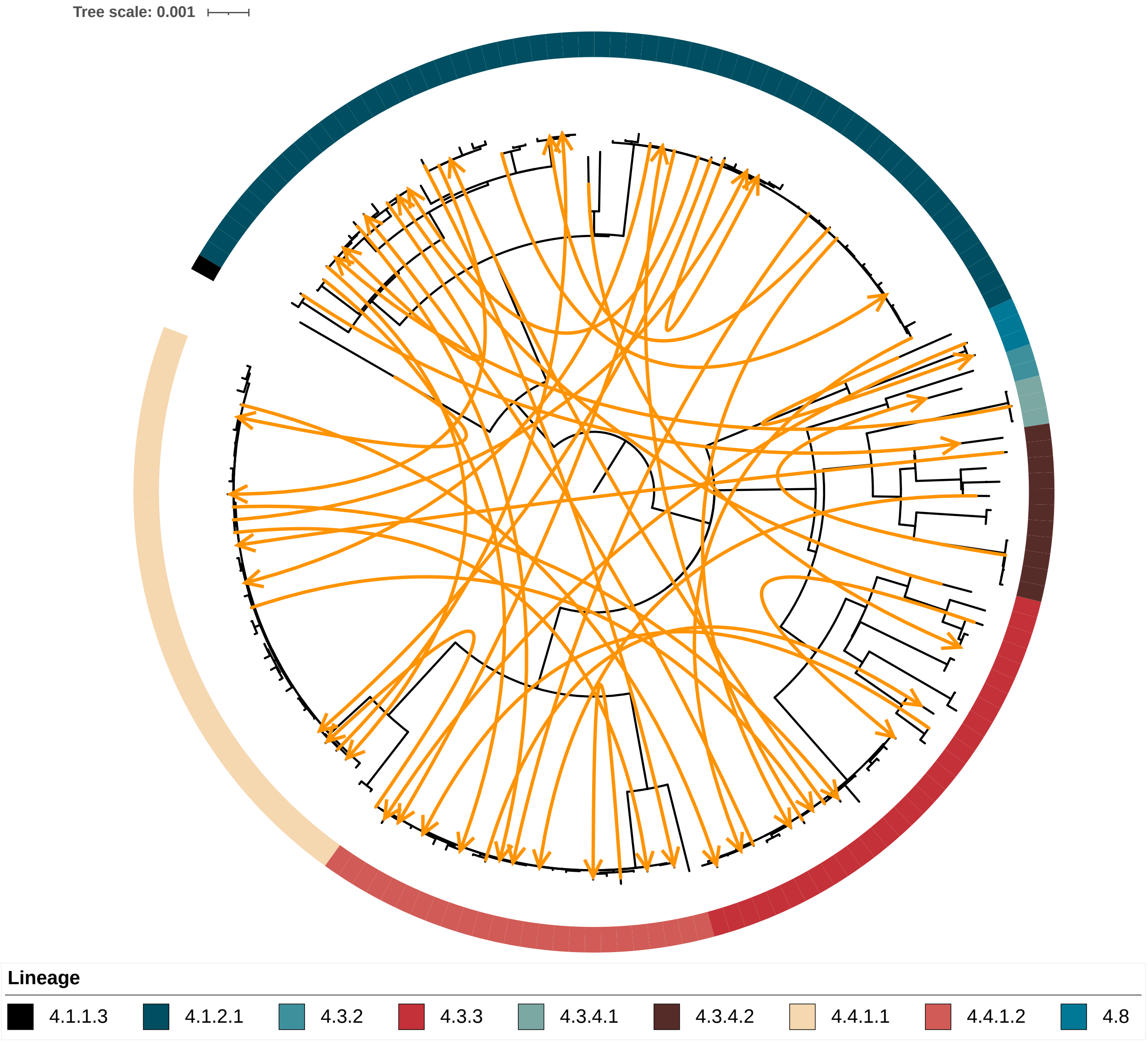


**Figure S1. Phylogenetic tree of *Mycobacterium tuberculosis* isolates from recurrent tuberculosis.** Branch lengths represent genetic distance in units of substitutions per site, and branches are colored according to sub-lineage classification. Cases of reinfection are depicted by orange connecting arrows that link pairs of isolates obtained from the same patient. The tree is scaled according to the substitution rate, as indicated by the scale bar located at the top left.
